# Clinical Relevance of Concomitant Large Artery Steno-occlusion and Risk of Recurrent Stroke in Atrial Fibrillation

**DOI:** 10.64898/2026.01.05.26343490

**Authors:** Hyung Seok Guk, Do Yeon Kim, Han-Gil Jeong, Jun Yup Kim, Beom Joon Kim, Moon-Ku Han, Jihoon Kang, Hyunsoo Kim, Kang-Ho Choi, Joon-Tae Kim, Kyu Sun Yum, Dong-Ick Shin, Dong-Seok Gwak, Dong-Eog Kim, Jong-Moo Park, Kyusik Kang, Jae Guk Kim, Soo Joo Lee, Minwoo Lee, Mi-Sun Oh, Kyung-Ho Yu, Byung-Chul Lee, Hong-Kyun Park, Yong-Jin Cho, Keun-Sik Hong, Joong-Goo Kim, Jay Chol Choi, Jeong-Ho Hong, Sung Il Sohn, Jin Kyo Choi, Tai Hwan Park, Jeong-Yoon Lee, Kyungbok Lee, Wook-Joo Kim, Jee-Hyun Kwon, Doo Hyuk Kwon, Jun Lee, Keon-Joo Lee, Wi-Sun Ryu, Ji Sung Lee, Juneyoung Lee, Philip B. Gorelick, Hee-Joon Bae, the CRCR-K-NIH investigators

## Abstract

**Background:** Recurrent ischemic stroke remains a major concern in patients with nonvalvular atrial fibrillation (NVAF) despite anticoagulation. However, not all NVAF-related strokes are purely cardioembolic—concomitant large artery steno-occlusion (cLASO) is frequently observed and may represent an independent contributor to stroke recurrence. Current guidelines address these cases but focusing almost exclusively on the cardioembolic source. This approach contrasts sharply with the management of acute coronary syndrome in patients with NVAF, where identifying and targeting the ’culprit lesion’ is standard practice. We evaluated whether the presence and clinical relevance of cLASO predict stroke recurrence in NVAF-related acute ischemic stroke (AIS).

**Methods:** This prospective multicenter cohort study enrolled 2,035 patients with NVAF-related AIS at 14 stroke centers in South Korea between October 2017 and April 2020. Patients underwent brain magnetic resonance imaging and angiography. cLASO, defined as any degree of stenosis or occlusion in major cerebral arteries, was categorized by anatomical severity (mild, moderate-to-severe, or occlusion) and clinical relevance. Clinically relevant cLASO was defined as a steno-occlusive lesion topographically concordant with the infarction, thereby sufficiently explaining the index stroke regardless of concurrent cardioembolic source; otherwise, lesions were classified as bystander. The primary outcome was recurrent ischemic stroke within 3 years, analyzed using competing risk analysis.

**Results:** Among 2,035 patients (mean age, 74.9 years; 54.8% male), 1,308 (64.3%) had cLASO, and 583 (28.6%) had clinically relevant cLASO. The 3-year cumulative incidence of recurrent ischemic stroke was 7.0%, with over 40% of recurrences occurring within the first month. Recurrence risk increased significantly with cLASO presence (4.5% vs. 8.1%), severity (mild, 5.7%; moderate-to-severe, 9.7%; occlusion, 9.1%) and clinical relevance (bystander, 3.8%; relevant, 13.9%) (all P’s < 0.05). In multivariable analysis, clinical relevance—rather than severity—was independently associated with recurrence (aHR, 4.10; 95% CI, 2.57-6.54).

**Conclusions:** Clinically relevant cLASO identifies a mechanistically distinct, high-risk phenotype that warrants a paradigm shift from a uniform cardioembolic model toward a ’lesion-specific approach. The early clustering of recurrence suggests an urgent window for intervention. This well-characterized phenotype may represent a potential target population for future trials evaluating intensified antithrombotic strategies that address both cardioembolic and atherothrombotic pathways.

**Clinical Perspective:** *What is New?:*

- Two-thirds of patients with AF-related stroke harbor concomitant large artery steno-occlusion (cLASO), yet current guidelines provide no recommendations for this common dual-mechanism scenario
- This prospective multicenter study introduces a novel classification distinguishing clinically relevant cLASO from bystander atherosclerosis; only clinical relevance—not anatomic severity—independently predicted stroke recurrence, conferring a fourfold increased risk.
- Over 40% of recurrences occurred within the first month, with relevant cLASO conferring a 6.0% early recurrence risk—substantially exceeding reported annual major hemorrhage risks in landmark anticoagulant trials.

*What Are the Clinical Implications?:*

- AF-related stroke is mechanistically heterogeneous; assuming all cases are cardioembolic may lead to suboptimal risk stratification and missed opportunities for targeted prevention.
- Routine vascular imaging should assess not merely stenosis severity, but topographic concordance with the infarct pattern—a straightforward approach using standard DWI and MRA that can be applied in diverse healthcare settings.
- The early clustering of recurrence in patients with relevant cLASO highlights a critical unmet need and identifies a well-characterized target population for future trials evaluating intensified antithrombotic strategies.

## Introduction

Prevention of recurrent ischemic stroke is a major clinical challenge in patients with nonvalvular atrial fibrillation (NVAF), even among those receiving anticoagulation.^1^ While oral anticoagulants effectively reduce cardioembolic stroke risk, recurrence rates remain considerable—approximately 4–7% at 1 year and up to 10% at 3 years.^1–3^ This persistent residual risk highlights a critical limitation in current management: a reductionist view that predominantly attributes NVAF-related strokes to a singular cardioembolic mechanism while overlooking alternative or concurrent vascular contributors.^4^

Cerebral atherosclerosis is frequently observed in patients with NVAF, sharing a common soil of vascular risk factors—such as hypertension, diabetes, and dyslipidemia—that promote both atrial cardiopathy and large artery disease. ^4–6^ Despite this overlap, current stroke prevention guidelines remain heavily cardio-centric, focusing almost exclusively on the cardioembolic source. ^1,7,8^ This approach contrasts sharply with the management of acute coronary syndrome in patients with NVAF, where identifying and targeting the ’culprit lesion’ is standard clinical practice.^7^ In the brain, however, the presence of concomitant large artery steno-occlusion (cLASO) is often dismissed as an incidental finding, and its prognostic significance—particularly according to its mechanistic contribution—remains largely unexplored ^9,10^.

A major barrier to refined risk stratification is the lack of a clear distinction between mechanistically relevant atherosclerosis and mere bystander findings. cLASO can be described by anatomic severity (mild, moderate-to-severe, or occlusion), but more crucially, by its clinical relevance—whether the lesion plausibly explains the index stroke independent of cardioembolism. To date, few studies have jointly applied diffusion-weighted imaging (DWI) and angiography to systematically evaluate these lesion characteristics in the context of NVAF.^11^ ^12^

We therefore aimed to determine whether cLASO, as identified by magnetic resonance angiography (MRA), is associated with an increased risk of recurrent ischemic stroke in patients with NVAF-related acute ischemic stroke (AIS). By challenging the uniform cardioembolic model and evaluating both anatomic severity and clinical relevance, we sought to identify a high-risk phenotype that warrants a more nuanced, lesion-specific approach to stroke prevention

## Methods

### Study Design and Participants

This was a prospective, multicenter cohort study including patients with NVAF-related AIS enrolled in the East Asian iSchemic sTroke with Atrial Fibrillation (EAST-AF) study, a substudy of the Clinical Research Collaboration for Stroke in Korea–National Institute of Health (CRCS-K-NIH) registry.^13–15^ Consecutive patients admitted to 14 tertiary stroke centers in South Korea between October 2017 and April 2020 were screened. NVAF was defined as documented atrial fibrillation in the absence of moderate-to-severe mitral stenosis or a mechanical prosthetic heart valve.

Patients were eligible if they had AIS confirmed by DWI and underwent MRA during the index hospitalization. Those without MRA were excluded. The registry protocol, imaging acquisition, and follow-up procedures were standardized across centers. All participants or their legally authorized representatives provided written informed consent, and the study was approved by the institutional review boards of all participating centers (B-1705/396-306).

### Neuroimaging Acquisition and Assessment

Brain magnetic resonance imaging and MRA were performed during the index hospitalization using 1.5- or 3.0-T scanners, according to harmonized institutional protocols. Sequences included DWI, fluid-attenuated inversion recovery (FLAIR), T2-weighted imaging, and time-of-flight MRA of intracranial and extracranial vessels. When indicated, contrast-enhanced MRA, computed tomography angiography (CTA) or catheter angiography was additionally performed.

All imaging data were centrally reviewed by two board-certified vascular neurologists (HS Guk, DY Kim) blinded to clinical outcomes. Discrepancies were resolved by consensus. Interobserver agreement was excellent for all imaging parameters (weighted κ’s > 0.90). Inter-modality agreement between MRA and CTA was validated in a subset of patients who underwent both modalities within 1 day (weighted κ = 0.77) (Supplemental Methods 1-2).

### Definition and Characterization of cLASO

cLASO was defined as any stenosis or occlusion in major intracranial or extracranial arteries—including internal carotid, middle cerebral, anterior cerebral, posterior cerebral, basilar, and vertebral arteries—on MRA or CTA.^16^ Stenosis severity was measured using the Warfarin–Aspirin Symptomatic Intracranial Disease (WASID) method.^17^

Each lesion was characterized by anatomic severity (mild [1–50%], moderate-to-severe [51–99%], or occlusion [no antegrade flow]) and clinical relevance. In patients with concurrent AF, determining whether a steno-occlusive lesion contributed to the index stroke—or merely coexisted—poses a challenge. We therefore applied a parsimonious approach: a lesion was classified as clinically relevant cLASO only if it carried sufficient explanatory weight, based on lesion severity and topographic concordance with the infarct pattern on DWI^11^ to account for the index stroke independent of the cardioembolic source. Specifically, this included lesions in which all acute infarcts on DWI could be attributed to a single arterial lesion, or residual steno-occlusion identified after thrombectomy indicating an underlying atherosclerotic substrate.^18^ Lesions not meeting these criteria were classified as bystander. For patients with multiple lesions, the most severe lesion determined the severity category, and the presence of any relevant lesion defined clinical relevance.

### Clinical Data Collection

Demographic characteristics, vascular risk factors, stroke severity (National Institutes of Health Stroke Scale [NIHSS] score), CHA₂DS₂-VASc (congestive heart failure, hypertension, age, diabetes mellitus, history of stroke/transient ischemic attack, vascular disease including myocardial infarction or peripheral artery disease and sex) score,^19^ index stroke characteristics, and discharge medications were prospectively collected using standardized CRCS-K-NIH protocols.^13,14^ Antithrombotic and statin therapy were recorded at discharge and during follow-up, and NOAC dosing was classified as guideline-recommended or non-recommended based on the 2021 European Heart Rhythm Association guidelines.^20^

### Outcomes

The primary outcome was recurrent ischemic stroke within 3 years, defined as a new focal neurological deficit of vascular origin persisting for at least 24 hours or a new acute infarct on DWI.^21^ Secondary outcomes were all-cause mortality and a composite of recurrent stroke (ischemic or hemorrhagic), myocardial infarction, or death. Myocardial infarction was diagnosed based on typical ischemic symptoms accompanied by either electrocardiographic changes or elevations in acute cardiac biomarkers.^21^ Outcome events were captured at clinical visits or structured telephone follow-ups at 3 months, 1, 2, and 3 years post-stroke.^13–15^

### Statistical Analysis

Patients with missing information on major covariates were excluded from the analysis (<0.2% of the total cohort). To address potential selection bias arising from exclusion of patients without DWI or MRA, we compared baseline characteristics between included and excluded patients. Baseline characteristics were compared using the χ² test for categorical variables and Mann–Whitney U test for continuous variables. The cumulative incidence of recurrent ischemic stroke was estimated using Fine–Gray competing risk models treating all-cause death as a competing event; all-cause death and the composite outcome were analyzed using the Kaplan–Meier method, with group differences assessed using Gray or log-rank tests.

Multivariable analyses employed Fine–Gray subdistribution hazard models for recurrent ischemic stroke and marginal Cox proportional hazards models for mortality and composite outcomes. Two strategies were applied: (1) cLASO presence vs. absence, and (2) joint modeling of cLASO severity and clinical relevance. In the latter model, patients without cLASO or with bystander cLASO served as a reference. Covariates were selected a priori, including age, sex, CHA₂DS₂-VASc components, smoking, discharge medications (antiplatelet, anticoagulant, statin), and discharge modified Rankin Scale (mRS). The proportional hazards assumption was verified before model fitting.

Sensitivity analyses were performed by excluding patients with complete occlusion to minimize misclassification of embolic occlusion as cLASO. A post hoc 1-month landmark analysis compared early (≤1 month) and late (>1 month to 3 years) recurrences. All statistical analyses were performed using SAS v9.4 (SAS Institute), with two-sided p <.05 considered significant. The study was conducted and reported according to the STROBE guidelines.^22^

## Results

### Patients Characteristics

Among 15,452 patients with AIS admitted to the participating centers during the study period, 2,563 were enrolled in the EAST-AF study, and 2,035 who underwent both DWI and MRA were included in this analysis (**Fig S1**). Patients without DWI or MRA were older and had more severe strokes, although vascular risk factor profiles were comparable **(Table S1).**

The mean age of included participants was 74.9 years, and 54.8% were male. The median NIHSS was 6, and the median CHA_2_DS_2_-VASc was 5 (**Table 1**). At discharge, 85% received anticoagulants (95% NOACs) and four-fifths of those on NOACs received a guideline-recommended dose. Anticoagulant prescription rates, particularly NOACs, remained stable throughout follow-up with apixaban being most frequently prescribed. Concurrent antiplatelet therapy was used in 14% of anticoagulated patients at discharge (Table S2).

**Table 1.** Baseline characteristics of patients with and without concomitant large artery steno-occlusion (cLASO)

| Characteristics | All<br>(n = 2035) | cLASO (-)<br>(n = 727) | cLASO (+)<br>(n = 1308) | P-value |
| --- | --- | --- | --- | --- |
| **Demographics & Stroke Characteristics** |  |  |  |  |
| Age, median (IQR) | 74.9±10.0 | 72.2±10.2 | 76.4±9.6 | <0.01 |
| Male, No. (%) | 1115 (54.8) | 458 (63.0) | 657 (50.2) | <0.01 |
| Time from symptom onset to admission (hours), median (IQR) | 2 (1 - 12) | 2 (1 - 11) | 3 (1 - 13) | 0.62 |
| Initial NIHSS, median (IQR) | 6 (2 - 14) | 5 (2 - 12) | 7 (3 - 15) | <0.01 |
| Pre stroke mRS, median (IQR) | 0 (0 - 1) | 0 (0 - 0) | 0 (0 - 1) | <0.01 |
| Discharge mRS, median (IQR) | 3 (1 - 4) | 2 (1 - 3) | 3 (2 - 5) | <0.01 |
| **Stroke Risk Factors** |  |  |  |  |
| Hypertension | 1477 (72.6) | 496 (68.2) | 981 (75.0) | <0.01 |
| Diabetes mellitus | 618 (30.4) | 187 (25.7) | 431 (33.0) | <0.01 |
| Dyslipidemia | 564 (27.7) | 175 (24.1) | 389 (29.7) | <0.01 |
| Stroke/TIA | 525 (25.8) | 150 (20.6) | 375 (28.7) | <0.01 |
| Coronary heart disease | 264 (13.0) | 76 (10.5) | 188 (14.4) | 0.01 |
| Peripheral artery disease | 47 (2.3) | 14 (1.9) | 33 (2.5) | 0.39 |
| Heart failure* | 75 (3.7) | 21 (2.9) | 54 (4.1) | 0.15 |
| Current smoking | 259 (12.7) | 120 (16.5) | 139 (10.6) | <0.01 |
| CHA<sub>2</sub>DS<sub>2</sub>-VASc score†, median (IQR) | 5 (4 - 6) | 5 (4 - 6) | 6 (5 - 6) | <0.01 |
| **Acute Recanalization Therapy** |  |  |  |  |
| IV thrombolysis | 390 (19.2) | 148 (20.4) | 242 (18.5) | 0.38 |
| Endovascular thrombectomy | 492 (24.2) | 208 (28.6) | 284 (21.7) | <0.01 |
| **Discharge medications** |  |  |  |  |
| Antithrombotics therapy |  |  |  | <0.01 |
| No antithrombotics | 122 (6.0) | 31 (4.3) | 91 (7.0) |  |
| Antiplatelet only | 183 (9.0) | 48 (6.6) | 135 (10.3) |  |
| Anticoagulant only | 1474 (72.4) | 596 (82.0) | 878 (67.1) |  |
| Combined | 256 (12.6) | 52 (7.2) | 204 (15.6) |  |
| Statin therapy | 1769 (86.9) | 636 (87.5) | 1133 (86.6) | 0.58 |

Data are presented as No. (%) unless otherwise indicated.
P values were derived from the $\chi^2$ test for categorical variables and the Mann–Whitney U test for continuous variables.
\* Congestive heart failure or dilated cardiomyopathy with ejection fraction < 40%
† CHA<sub>2</sub>DS<sub>2</sub>-VASc indicates congestive heart failure, hypertension, age ≥75 years, diabetes mellitus, prior stroke/TIA, vascular disease, age 65–74 years, and sex category (female).
Abbreviation: cLASO, concomitant large artery steno-occlusion; NIHSS, National Institutes of Health Stroke scale; mRS, modified Rankin Scale; TIA, transient ischemic attack; IQR, interquartile range.

### Distribution and Characteristics of cLASO

cLASO was present in 1,308 patients (64.3%), and 583 (44.6% of cLASO cases; 28.6% of the total cohort) had clinically relevant lesions **(Table 2, Table S3)**. By anatomic severity, 502 patients (38.4% of cLASO cases) had mild stenosis, 391(29.9%) moderate-to-severe stenosis, and 415 (31.7%) with occlusion. Clinically relevant lesions compared to bystander lesions were more often moderate-to-severe or occlusive, though 18.5% were mild stenosis (**Table S3**).

**Table 2.** Cumulative incidence of clinical outcomes according to the presence, severity, and clinical relevance of cLASO.

|  | Numbers<br>(Percent) | Recurrent Ischemic Stroke |  |  | All-cause death |  |  | Composite Event |  |  |
| --- | --- | --- | --- | --- | --- | --- | --- | --- | --- | --- |
|  |  | 1 month | 1 year | 3 years | 1 month | 1 year | 3 years | 1 month | 1 year | 3 years |
| **All Patients** | 2035 (100) | 2.9<br>(2.2-3.7) | 4.6<br>(3.8-5.6) | 7.0<br>(5.8-8.2) | 2.8<br>(2.1-3.5) | 13.3<br>(11.8-14.8) | 24.5<br>(22.4-26.6) | 6.0<br>(5.0-7.1) | 17.9<br>(16.2-19.6) | 30.4<br>(28.2-32.7) |
| **Absence** | 727 (35.7) | 1.2<br>(0.6-2.3) | 2.5<br>(1.5-4.0) | 4.5<br>(3.1-6.4) | 1.3<br>(0.5-2.2) | 8.2<br>(6.2-10.3) | 16.6<br>(13.3-19.9) | 3.1<br>(1.8-4.5) | 11.4<br>(9.0-13.8) | 20.6<br>(17.1-24.2) |
| **Presence** | 1308 (64.3) | 3.7<br>(2.8-4.8) | 5.7<br>(4.5-7.0) | 8.1<br>(6.7-9.8) | 3.5<br>(2.5-4.5) | 15.8<br>(13.8-17.7) | 28.4<br>(25.7-31.0) | 7.5<br>(6.1-8.9) | 21.1<br>(19.0-23.3) | 35.2<br>(32.4-38.0) |
| **P-value** |  | <.01 |  |  | <.01 |  |  | <.01 |  |  |
| **Severity** |  |  |  |  |  |  |  |  |  |  |
| Mild | 502 (38.4) | 1.8<br>(0.8-3.3) | 3.3<br>(1.9-5.2) | 5.7<br>(3.7-8.3) | 0.9<br>(0.0-1.7) | 9.9<br>(7.2-12.7) | 20.2<br>(16.1-24.2) | 2.9<br>(1.3-4.4) | 13.9<br>(10.7-17.0) | 26.9<br>(22.4-31.4) |
| Moderate-to-Severe | 391 (29.9) | 4.9<br>(3.0-7.5) | 6.6<br>(4.3-9.4) | 9.7<br>(6.8-13.2) | 2.2<br>(0.7-3.7) | 16.5<br>(12.7-20.3) | 30.2<br>(24.9-35.5) | 7.1<br>(4.5-9.8) | 22.3<br>(18.0-26.6) | 35.4<br>(30.1-40.8) |
| Occlusion | 415 (31.7) | 4.6<br>(3.0-6.6) | 7.0<br>(5.1-9.4) | 9.1<br>(6.7-11.9) | 6.6<br>(4.5-8.7) | 20.2<br>(16.8-23.6) | 34.1<br>(29.7-38.5) | 11.6<br>(8.9-14.2) | 26.4<br>(22.7-30.2) | 42.0<br>(37.4-46.6) |
| **P-value** |  | <.01 |  |  | <.01 |  |  | <.01 |  |  |
| **Relevance** |  |  |  |  |  |  |  |  |  |  |
| Bystander | 725 (55.4) | 2.1<br>(1.2-3.2) | 2.6<br>(1.6-3.9) | 3.8<br>(2.6-5.5) | 2.8<br>(1.7-4.0) | 13.8<br>(11.3-16.2) | 25.1<br>(21.8-28.5) | 5.1<br>(3.6-6.7) | 16.7<br>(14.1-19.4) | 28.9<br>(25.4-32.4) |
| Relevant | 583 (44.6) | 6.0<br>(4.3-8.1) | 9.8<br>(7.5-12.4) | 13.9<br>(11.1-17.0) | 4.4<br>(2.8-6.1) | 18.4<br>(15.3-21.6) | 32.7<br>(28.5-37.0) | 10.6<br>(8.1-13.1) | 27.0<br>(23.4-30.6) | 43.7<br>(39.3-48.2) |
| **P-value** |  | <.01 |  |  | <.01 |  |  | <.01 |  |  |

Values are presented as percentage (95% Confidence Interval). P values were calculated using Gray's test for recurrent ischemic stroke and the log-rank test for all-cause death and composite events.
Recurrent ischemic stroke was estimated using the cumulative incidence function considering all-cause death as a competing risk.
All-cause death and the composite outcome (recurrent stroke, myocardial infarction, or death) were estimated using the Kaplan–Meier method.
Abbreviation: cLASO, concomitant large artery steno-occlusion.

Steno-occlusion most frequently involved in the middle cerebral artery, followed by posterior and anterior cerebral arteries **(Table S4, Fig S2).** Participants with cLASO were older and more often female, and had higher vascular risk factor burdens, resulting in a one-point higher median CHA_2_DS_2_-VASc score (**Table 1**). At discharge, combination antithrombotic therapy was more than twice as common among patients with cLASO than those without (15.7% vs. 7.2%), most pronounced in patients with relevant cLASO (16.6%)—suggesting clinical recognition of concurrent atherosclerotic disease in this subgroup.

### Recurrent Ischemic Stroke

During a median follow-up of 756 days (IQR, 718-1096 days), 144 recurrent ischemic strokes occurred. Additionally, there were 11 hemorrhagic strokes, 446 deaths, and 31 myocardial infarctions. The 3-year cumulative incidence of recurrent ischemic stroke was 7.0%, with more than 40% of recurrences occurring within the first month (**Table 2**).

Recurrence increased with cLASO presence, anatomic severity and clinical relevance. **(Table 2**, **Fig 1)** Patients with cLASO had higher recurrence than those without (8.1% vs. 4.5% at 3 years; absolute difference, 3.6%). However, the pattern of recurrence risk differed markedly depending on how cLASO was characterized. When stratified by anatomic severity alone, a 3-year recurrence risk increased modestly; 5.7% for mild, 9.7% for moderate-to-severe and 9.1% for occlusion. By contrast, stratification by clinical relevance revealed a clear separation: patients with relevant cLASO had a 3-year recurrence of 13.9%, compared to 3.8% for bystander and 4.5% for those without cLASO. When both severity and relevance were considered, relevant cLASO consistently conferred higher recurrence risks across all severity categories, with the most pronounced effect observed in patients with moderate-to-severe stenosis (**Figure 2, Table S6**). Notably, bystander cLASO conferred no excess risk over absence of cLASO, underscoring that the mere presence of atherosclerosis without a mechanistic contribution does not elevate recurrence risk.

**Figure 1.**
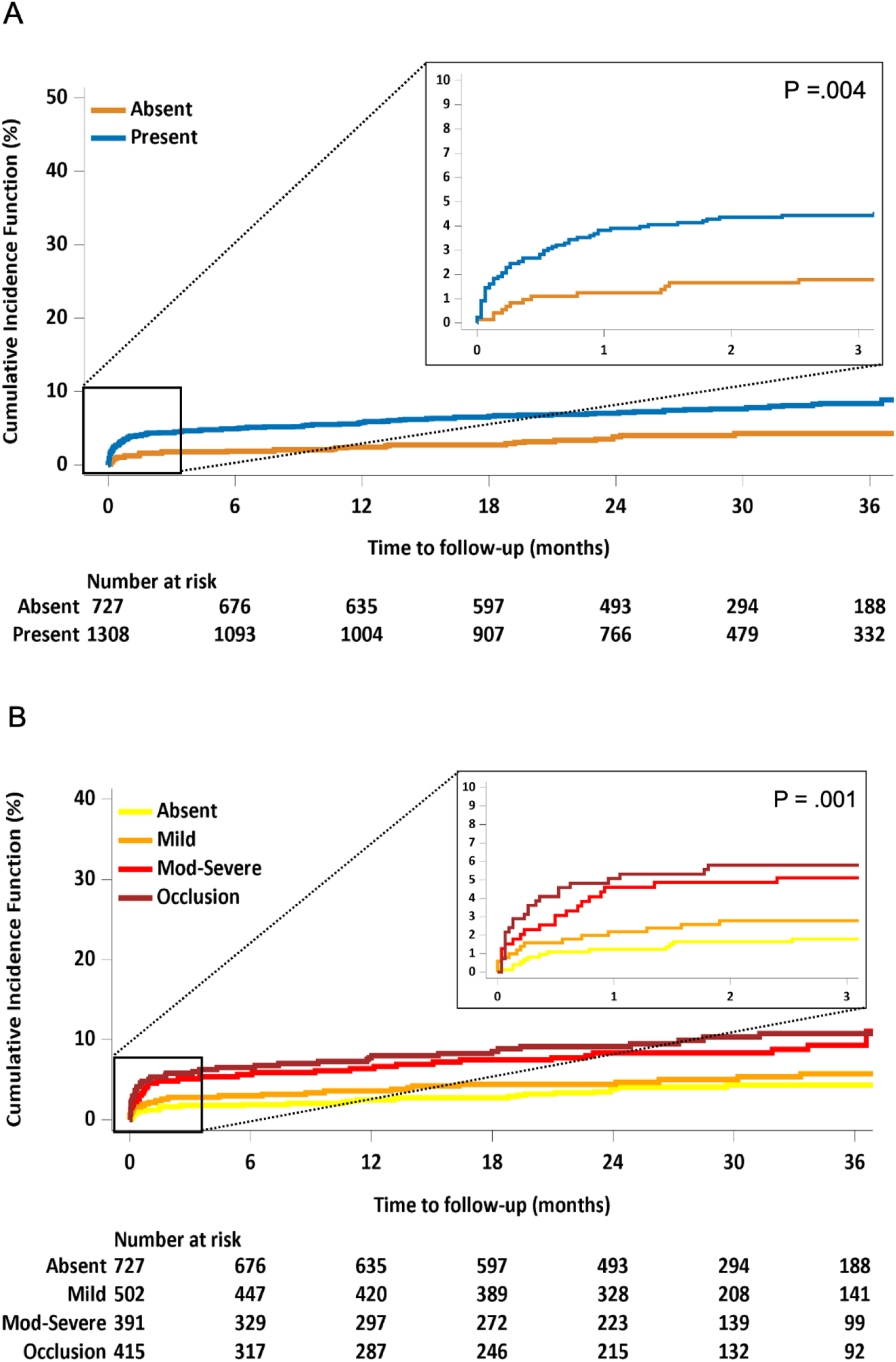
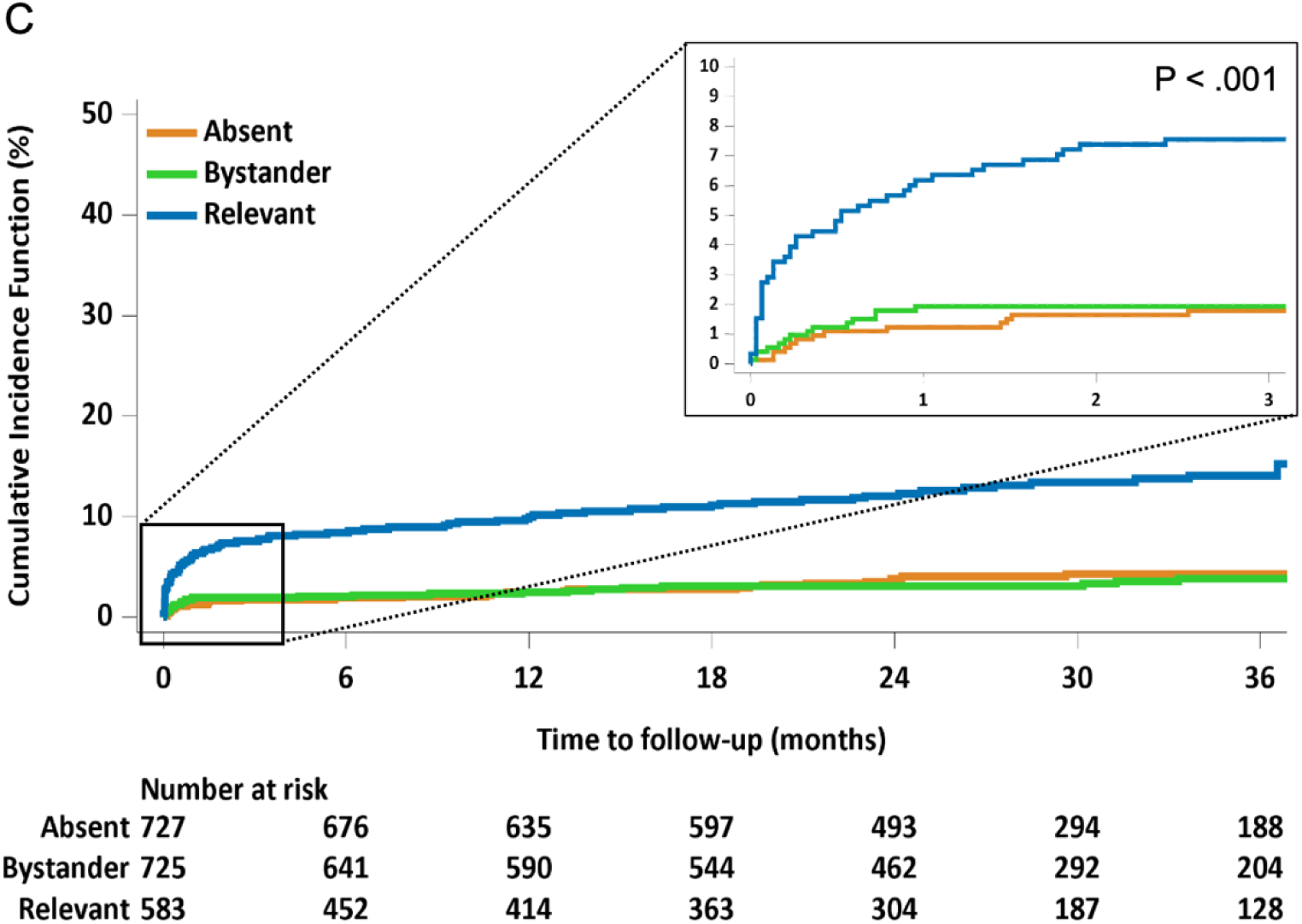
Cumulative incidence of recurrent ischemic stroke according to cLASO presence, anatomic severity, and clinical relevance. Cumulative incidence curves displaying time to recurrent ischemic stroke by cLASO presence (A), anatomic severity (B) and clinical relevance (C). Cumulative incidence curves from Fine–Gray competing risk analysis with all-cause death as the competing event. Numbers at risk are from Kaplan–Meier estimates. The insets display the cumulative incidence during the first 3 months post-stroke. P values were calculated using Gray’s test. Abbreviation: cLASO, concomitant large artery steno-occlusion.

**Figure 2.**
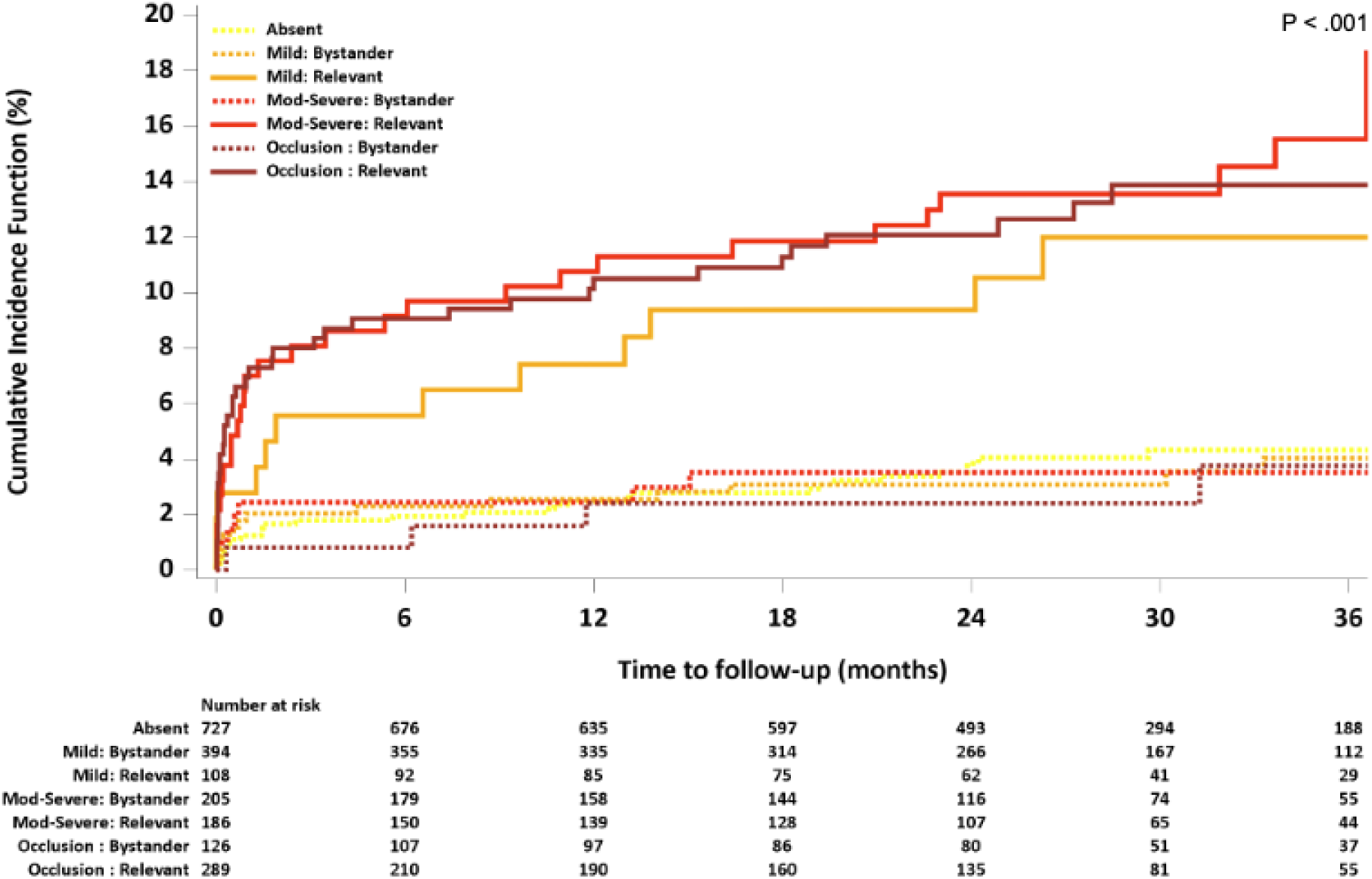
Cumulative incidence of recurrent ischemic stroke stratified by both anatomic severity and clinical relevance of cLASO. Cumulative incidence curves from Fine–Gray competing risk analysis with all-cause death as the competing event. Patients were stratified by both anatomic severity (no, mild, moderate-to-severe, occlusion) and clinical relevance (no, bystander, relevant). Across all severity categories, relevant cLASO (solid lines) consistently conferred higher recurrence risk than bystander lesions (dashed lines) of the same severity. Bystander lesions, regardless of severity, showed recurrence rates similar to patients without cLASO. Abbreviation: cLASO, concomitant large artery steno-occlusion.

The excess risk associated with relevant cLASO was evident early, with a 1-month cumulative incidence of 6.0% versus 1.2% in patients without cLASO (absolute risk difference of 4.8%). In multivariable Fine–Gray models, a fundamental decoupling of severity and relevance was observed. While cLASO presence was associated with nearly doubled recurrence risk (aHR 1.97; 95% CI, 1.60–2.42), when severity and clinical relevance were modeled together, only clinical relevance remained a selective and potent predictor of recurrence (aHR, 4.10; 95% CI, 2.57–6.54); anatomic severity showed no independent association (**Table 3**). This fourfold increased risk persisted despite more frequent use of combination antithrombotic therapy in patients with relevant cLASO.

**Table 3.** Multivariable analysis of clinical outcomes according to cLASO features.

|  | Recurrent Ischemic Stroke |  |  |  | All-cause death |  |  |  | Composite Event |  |  |  |
| --- | --- | --- | --- | --- | --- | --- | --- | --- | --- | --- | --- | --- |
|  | Model 1 |  | Model 2 |  | Model 1 |  | Model 2 |  | Model 1 |  | Model 2 |  |
|  | HR<br>(95% CI) | P-value | HR<br>(95% CI) | P-value | HR<br>(95% CI) | P value | HR<br>(95% CI) | P-value | HR<br>(95% CI) | P value | HR<br>(95% CI) | P-value |
| **Presence** |  |  |  |  |  |  |  |  |  |  |  |  |
| Absence | 1 (Ref) |  |  |  | 1(Ref) |  |  |  | 1(Ref) |  |  |  |
| Presence | 1.97<br>(1.60-2.42) | <.001 |  |  | 1.30<br>(1.00-1.69) | 0.048 |  |  | 1.42<br>(1.13-1.79) | <0.01 |  |  |
| **Severity** |  |  |  |  |  |  |  |  |  |  |  |  |
| Absence |  |  | 1(Ref) |  |  |  | 1(Ref) |  |  |  | 1(Ref) |  |
| Mild |  |  | 0.78<br>(0.55-1.11) | 0.93 |  |  | 0.97<br>(0.73-1.29) | 0.84 |  |  | 1.02<br>(0.80-1.30) | 0.87 |
| Moderate-to-severe |  |  | 0.98<br>(0.59-1.61) | 0.82 |  |  | 1.42<br>(1.00-2.02) | 0.049 |  |  | 1.30<br>(0.89-1.90) | 0.17 |
| Occlusion |  |  | 0.93<br>(0.51-1.70) | 0.17 |  |  | 1.40<br>(0.94-2.08) | 0.10 |  |  | 1.32<br>(0.91-1.92) | 0.15 |
| **Relevance** |  |  |  |  |  |  |  |  |  |  |  |  |
| Absence |  |  | 1(Ref) |  |  |  | 1(Ref) |  |  |  | 1(Ref) |  |
| Bystander |  |  |  |  |  |  |  |  |  |  |  |  |
| Relevant |  |  | 4.10<br>(2.57-6.54) | <.001 |  |  | 1.10<br>(0.84-1.45) | 0.48 |  |  | 1.43<br>(1.25-1.64) | <.01 |

Hazard ratios (HRs) are from Fine–Gray subdistribution hazard models for recurrent ischemic stroke (all-cause death as a competing risk) and from marginal Cox proportional hazards models for all-cause death and the composite outcome. Model 1 examines cLASO presence; Model 2 examines severity and relevance in the same model. Adjusted covariates include age, sex, CHA<sub>2</sub>DS<sub>2</sub>-VASc score components, smoking status, discharge medications (antiplatelets, anticoagulants, and statins), and discharge modified Rankin Scale (mRS) score (0–1 vs. ≥2).
Abbreviation: Ref indicates reference; HR, hazard ratio; CI, confidence interval.

### Secondary Outcomes

The 3-year cumulative incidence of death (24.5%) was more than three times higher than that of recurrent ischemic stroke (7.0%). Particularly, the predictive patterns for mortality diverged fundamentally from those observed for stroke recurrence. Mortality was higher in patients with cLASO than those without (28.4% vs 16.6%, P < .01) (**Table 2, Fig S3**). Unlike the stroke recurrence, which was driven by mechanistic relevance, mortality was more closely aligned with anatomic severity—acting as a surrogate for systemic vascular burden. The risk of death increased monotonically with severity: 16.6% without cLASO, 20.2% with mild stenosis, 30.2% with moderate-to-severe stenosis, and 34.1% with occlusion (P for trend <.01). By clinical relevance, mortality was 25.1% for bystander and 32.7% for clinically relevant lesions (P < .01). In multivariable models including both severity and relevance, this divergence was confirmed: anatomic severity (moderate-to-severe stenosis) retained its association with mortality (aHR, 1.42; 95% CI, 1.00–2.02), whereas clinical relevance was not independently associated with death (**Table 3 and S5, Fig S4**).

The patterns of composite outcomes (stroke, myocardial infarction, or death) were largely consonant with those observed for mortality (**Table 2, Table S5, Fig S5**). In the integrated model, clinical relevance showed a modest association (adjusted HR 1.43; 95% CI 1.25–1.64), while severity was not significant (**Table 3**).

### Sensitivity and Post-hoc Analyses

Sensitivity Analysis excluding patients with occlusion yielded consistent results, with clinical relevance remaining strongly associated with recurrence (aHR, 3.88; 95% CI, 2.17–5.56) independent of severity (**Table S7**). In landmark analysis, the excess risk associated with relevant cLASO persisted beyond the early high-risk period; hazard ratios for relevant cLASO was similar for early (within 1 month, adjusted hazard ratio [aHR], 4.09; 95% confidence interval [CI], 2.70-6.19) and late (after 1 month, aHR 3.99; 95% CI, 2.69-5.92) recurrence (**Fig 3, Table S8**), confirming that relevant cLASO identifies a high-risk phenotype that confers both immediate mechanistic and sustained long-term risk.

**Figure 3.**
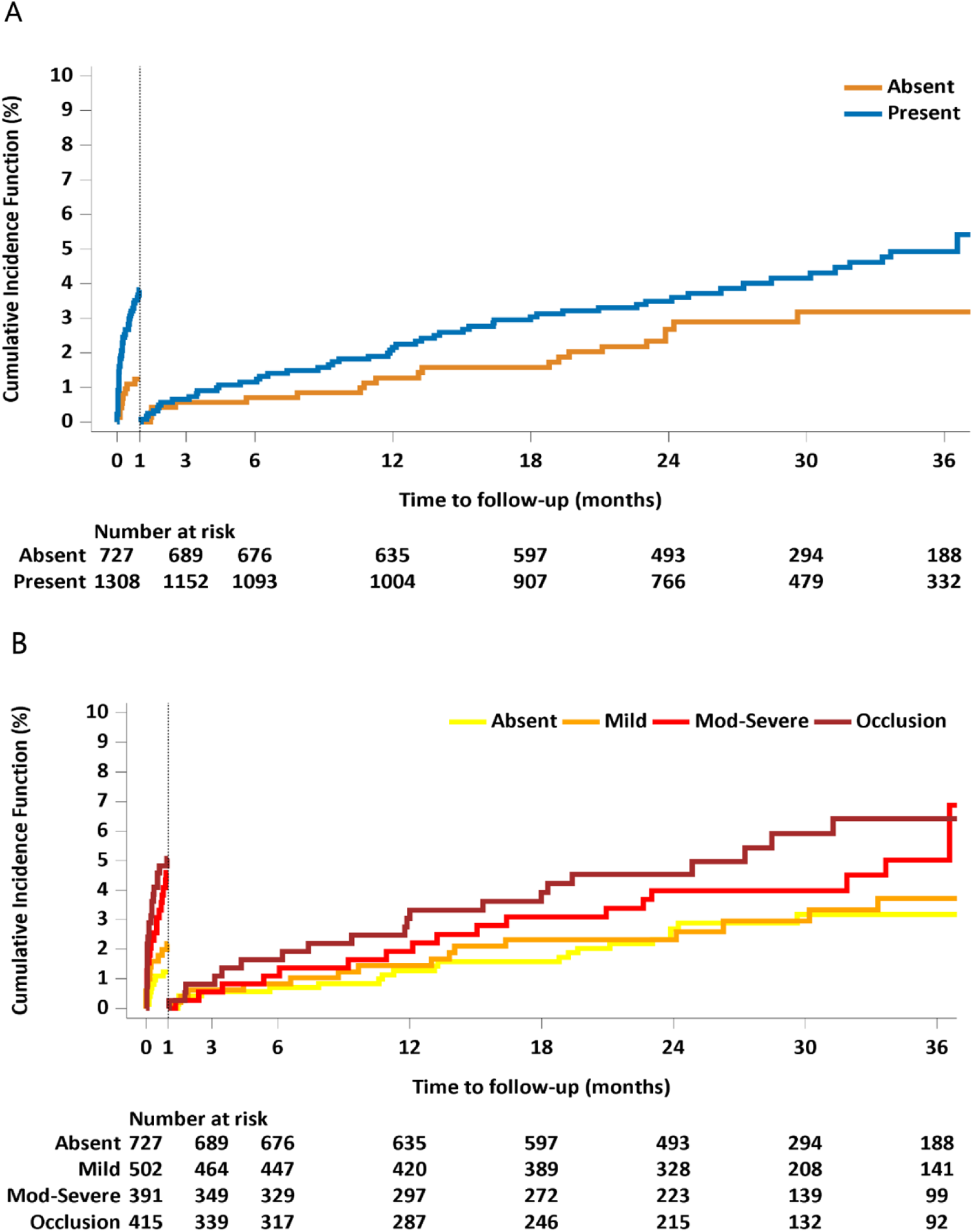
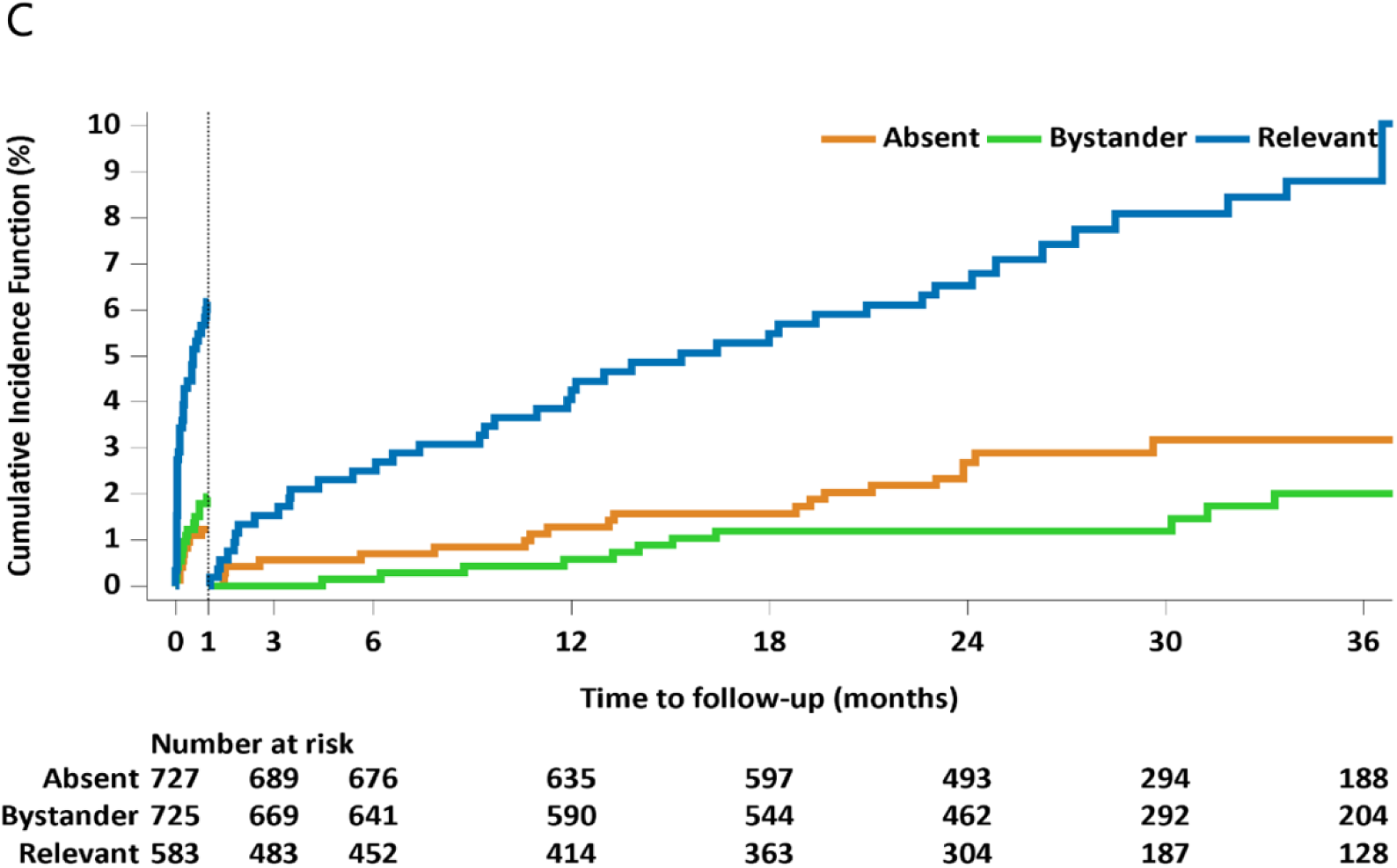
Landmark analysis of recurrent ischemic stroke at the 1-month time point. Cumulative incidence curves from Fine–Gray competing risk analysis beginning 1 month after index stroke, with all-cause death as the competing event. Patients were stratified by cLASO presence (A), anatomic severity (B), and clinical relevance (C). P values from Gray’s test are provided for the periods within and after 1 month separately. All three stratifications were significantly associated with recurrence in both time periods (all P<0.01). Clinical relevance showed the widest separation in cumulative incidence between groups. Abbreviation: cLASO, concomitant large artery steno-occlusion.

## Discussion

In this multicenter prospective cohort of patients with NVAF-related AIS, cLASO was identified in nearly two-thirds of the cohort, and nearly two-fifth of these lesions were clinically relevant to the index stroke. This high prevalence raises the possibility that stroke mechanisms in patients with AF may be more heterogeneous than traditionally assumed. The presence of cLASO was independently associated with increased risks of recurrent ischemic stroke, mortality, and composite outcomes. When anatomic severity and clinical relevance were modeled together, only clinically relevant cLASO was independently associated with recurrent stroke, conferring a more than fourfold increased risk. Patients with bystander cLASO, by contrast, had recurrence rates comparable to those without cLASO, suggesting that the mere coexistence of atherosclerosis does not confer additional risk. These observations highlight the potential importance of distinguishing mechanistically contributory lesions from incidental findings in this population. Importantly, this distinction was reproducible, with excellent inter-rater agreement, suggesting that clinical relevance can be reliably assessed using routine neurovascular imaging.

The distinction between relevant and bystander cLASO may have implications for both clinical practice and trial design. Current treatment algorithms, informed by the TOAST classification, recommend anticoagulation for cardioembolic stroke but lack specific guidance for concurrent large artery atherosclerosis.^1,7^ Our data demonstrate that this lack of guidance creates a significant unmet need: patients with clinically relevant cLASO faced a staggering 6.0% recurrence risk within the first month—a five-fold increase compared to those without cLASO (1.2%). This absolute excess risk of 4.8% within only 30 days substantially exceeds the annual major hemorrhage risk reported in landmark anticoagulant trials. ^24,28,29^ Such a disparity proves that the current cardio-centric approach leaves this dual-mechanism phenotype dangerously under-protected during their most vulnerable period. Particularly, this excess risk persisted despite more frequent use of combination antithrombotic therapy in these patients, suggesting that current treatment adjustments may be insufficient.

Beyond the magnitude of risk, the temporal clustering of events provides a clear target for intervention. More than 40% of recurrences occurred within the first month, with a particularly marked early hazard among those with moderate-to-severe stenosis. These patterns suggest that effective intervention must be initiated within this critical early hazard window. Specifically, we propose that future trials evaluate an intensified, ’front-loaded’ antithrombotic strategy—such as the addition of short-term antiplatelet therapy to full-dose anticoagulation during the initial 30 days post-stroke—to bridge the gap where anticoagulation alone is insufficient. While landmark analysis demonstrated that this excess risk persists throughout follow-up, the acute phase represents the most urgent window for intervention.

A recent trial evaluated adding antiplatelet therapy to NOACs in AF patients with systemic atherosclerosis but found no benefit.^30^ However, randomization occurred at a median of 3 weeks, likely missing the highest risk window identified in our study. Furthermore, by defining atherosclerosis using systemic rather than cerebrovascular criteria, the trial may have included bystander lesions that dilute treatment effects. Our findings extend those of prior studies by demonstrating that clinically relevant cLASO identified acutely carries a distinct, sustained recurrence risk that necessitates early, precisely targeted intervention.

The risk profile of recurrent stroke in NVAF-related AIS appears to be evolving. While cardioembolic stroke was historically thought to carry higher recurrence risk than atherosclerotic subtypes,^31–33^ the gap has markedly diminished in the NOAC era. ^28,34,35^ This shift likely reflects the effectiveness of modern anticoagulation in reducing embolic recurrences.^36–38^ To achieve further reductions, identifying non-embolic contributors like atherothrombosis—which involves both platelet-mediated initiation and coagulation-driven propagation—is essential. Our findings demonstrate that for patients with relevant cLASO, NOAC therapy alone may be insufficient, and novel antithrombotic strategies warrant investigation.

Our findings also highlight the prognostic significance of clinically relevant mild (≤50%) stenosis. In our cohort, these patients had a 3-year recurrence risk of 11.8%, suggesting they may also derive benefit from intensified secondary prevention. This aligns with prior studies, such as TIAregistry.org, showing that even mild atherosclerosis increases vascular risk.^39^ The prevalence and severity distribution of cLASO in our study are consistent with findings from OXVASC^16^ and CICAS^40^, reinforcing the importance of comprehensive vascular imaging to identify both severe and clinically relevant mild lesions.

This study has limitations. First, only 79% of patients underwent MRA, potentially introducing selection bias. However, the CHA₂DS₂-VASc and risk profiles were similar to other AF populations.^36,41^ Second, MRA may overestimate stenosis,^42^ but our validation against CTA showed substantial agreement. Third, we did not stratify by AF subtype. which may potentially affect risk of recurrent stroke.^43^ Fourth, although conducted in a South Korean population, the global relevance of our findings is supported by the rising burden of large artery disease in Western populations.^44–45^ Moreover, our assessment of ’clinical relevance’—based on topographic concordance between DWI and angiography—represents a universal, race-agnostic methodology that is readily applicable in any setting with standard neuroimaging. Finally, as an observational study, residual confounding cannot be excluded.

In summary, in the contemporary NOAC era, clinically relevant cLASO emerges as a reproducibly identifiable, high-risk phenotype. These patients exhibit a distinct recurrence pattern—early onset with sustained long-term risk—that persists despite current anticoagulation strategies. Our findings underscore the need for randomized trials evaluating intensified or novel antithrombotic approaches in this well-defined subgroup, with early initiation and precise patient selection based on clinical relevance.

## Data Availability

The data that support the findings of this study are available from the corresponding author on reasonable request and approval by the CRCS-K-NIH investigators.

## Acknowledgement

The authors thank all research collaborators, research nurses and participating patients in the different centers participating in the EAST-AF cohorts and CRCS-K-NIH collaborations.

## Authors

Hyung Seok Guk (Seoul National University Bundang Hospital)

Do Yeon Kim (Seoul National University College of Medicine, Seoul National University Bundang Hospital)

Han-Gil Jeong (Seoul National University Bundang Hospital)

Jun Yup Kim (Seoul National University Bundang Hospital, Seoul National University College of Medicine)

Beom Joon Kim (Seoul National University Bundang Hospital Department of Neurology)

Moon-Ku Han (epartment of Neurology, Seoul National University Bundang Hospital)

Jihoon Kang (Seoul National University Bundang Hospital)

Hyunsoo Kim (Chonnam National University Hospital)

Kang-Ho Choi (Chonnam National University Hospital)

Joon-Tae Kim (Chonnam National University Hospital)

Kyu Sun Yum (Chungbuk National University College of Medicine)

Dong-Ick Shin (Chungbuk National University Hospital)

Dong-Seok Gwak (Dongguk University Ilsan Hospital) Dong-Eog Kim (Dongguk University Ilsan Hospital)

Jong-Moo Park (Uijeongbu Eulji Medical Center, Eulji University)

Kyusik Kang (Nowon Eulji Medical Center, Eulji University)

Jae Guk Kim (Daejeon Eulji Medical Center, Eulji University School of Medicine)

Soo Joo Lee (Eulji University Hospital)

Minwoo Lee (Hallym University Sacred Heart Hospital)

Mi Sun Oh (Hallym University Sacred Heart Hospital, University College of Medicine)

Kyung-Ho Yu (Hallym University Sacred Heart Hospital)

Byung-Chul Lee (Hallym University Sacred Heart Hospital, Hallym University College of Medicine)

Hong-Kyun Park (Inje University Ilsan Paik Hospital)

Yong-Jin Cho (Ilsan Paik Hospital, Inje University)

Keun-Sik Hong (Ilsan Paik Hospital, Inje University)

Joong-Goo Kim (Jeju National University Hospital)

Jay Chol Choi (Jeju National University)

Jeong-Ho Hong (Keimyung University Dongsan Hospital)

Sung-Il Sohn (Dongsan Hospital, Keimyung University School of Medicine)

Jin Kyo Choi (Seoul Medical Center)

Tai Hwan Park (Seoul Medical Center)

Jeong-Yoon Lee (Soonchunhyang University Hospital)

Kyungbok Lee (Soonchunhyang University College of Medicine)

Wook-Joo Kim (Ulsan University Hospital, University of Ulsan College of Medicine)

Jee-Hyun Kwon (Ulsan University Hospital Department of Neurology)

Doo Hyuk Kwon (Yeungnam University Medical Center)

Jun Lee (Yeungnam University Medical Center, Daegu, Republic of Korea)

Keon-Joo Lee (Korea University Guro Hospital)

Wi-Sun Ryu (JLK Inc)

Ji Sung Lee (Asan Medical Center)

Juneyoung Lee (Korea University College of Medicine)

Philip Gorelick (Northwestern University)

Hee-Joon Bae (Cerebrovascular Center, Seoul National University Bundang Hospital)

